# Assessment of relationship between Google Trend search data on clinical symptoms and cases reported during the first wave of COVID-19 outbreak in India

**DOI:** 10.1101/2023.06.08.23291183

**Authors:** Hariprasad Vavilala, Rajasekhar Mopuri, Srinivasa Rao Mutheneni

**Author notes:** **Corresponding Author:** Dr M Srinivasa Rao Principal Scientist Coordinator EIACP Resource Partner of Climate Change & Public Health Applied Biology Division, CSIR-Indian Institute of Chemical Technology Tarnaka, Hyderabad-500007, Telangana, India. ORCID: https://orcid.org/0000-0003-3263-3905.

## Abstract

Infodemiology and infoveillance approaches have been extensively used in recent years to support traditional epidemiology and disease surveillance. Hence, the present study aimed to explore the association between Google Trends (GTs) search of clinical symptoms and cases reported during the first wave of COVID-19. The GT data from January 30, 2020, to September 30, 2020, and daily COVID-19 cases in India and a few selected states were collected from the Ministry of Health and Family Welfare, Government of India. Correlation analysis was performed between the GT index values and the number of confirmed cases. Followed by, the COVID-19 cases were predicted using Bayesian regression and classical linear regression models. A strong association was observed between the search index of clinical symptoms and reported COVID-19 cases (cold: R=0.41, headache: R=0.46, fever: R=0.66, loss of taste: R=0.78, loss of smell R=0.86) across India. Similarly, lagged correlations were also observed (loss of smell, loss of taste, loss of taste and loss of smell, fever and headache show 3, 9, 1, 9, and 13 days lag periods respectively). Besides this, the Bayesian regression model was outperformed (MAE: 0.331164, RMSE: 0.411087) for predicting the COVID-19 cases in India and regionally than the frequentist linear regression (MAE: 0.33134, RMSE: 0.411316). The study helps health authorities better prepare and planning of health care facility timely to avoid adverse impacts.

## 1. Introduction

The coronavirus disease 2019 (COVID-19) caused by the severe acute respiratory syndrome coronavirus-2 (SARS-CoV-2) is a bat-derived virus. It is a *betacoronavirus* of the family *Coronaviridae* with <140 nm in size [1,2]. The other coronaviruses in this family are HCoV-229E, -HKU1, -NL63 and -OC43, which are circulating among humans and causing mild respiratory illness [3]. Since its emerging in Wuhan, China in late 2019, the COVID-19 has spread rapidly around the globe and established local transmission [1,4]. On January 30, 2020, World Health Organisation (WHO) declared COVID-19 outbreak as a public health emergency of international concern and on March 11, 2020, notified as a pandemic [5]. As of November 18, 2022, there have been 633,601,048 confirmed cases of COVID-19, including 6,596,542 deaths observed across the globe [6]. The SARS-CoV-2 is a single-stranded RNA virus consist of 29,903 bp and it targets the angiotensin-converting enzyme (ACE)-2 receptors to enter into the host cells [7]. Due to the unavailability of specific medicines or vaccines, insufficient evidence regarding the transmission mechanism of the disease also make it difficult to fight against the disease. Hence, many countries have imposed nationwide lockdown, followed by social distancing, sanitization, facemasks made compulsory and non-pharmaceutical interventions (NPIs) particularly "trace, quarantine, test, isolate and treat" made a great contribution towards controlling the pandemic [8,9].

Internet availability and internet users have grown rapidly in recent years similarly, the health-related information on the internet has also changed how people seek information about health [10]. An internet-based novel surveillance system supported by the internet search behaviour of the community has recently increased and emerging as a promising technique in epidemiology and public health. Data generated from queries fed into search engines recorded and used for surveillance to understand the trend of the diseases [10]. Large volumes of aggregated big data on an internet search are available at population scale and in near real time where the internet users are adequate [11]. The organisations like Centre for Disease Control and Prevention (CDC) has already been using this internet and social media data as an intervention tool for sexually transmitted diseases. However, these internet technologies used as a supplement for existing technologies to mitigate the shortcomings of health monitoring [12].

In recent years, Infodemiology (epidemiology) and infoveillance (surveillance) are an integral part of public health informatics research using web-based data to analyse search behaviour on the internet. Infodemiology defined as the "science of distribution and determinants of information in an electronic medium, specifically the internet, or in a population, with the ultimate aim to inform public health and public policy". Similarly, the primary aim of infoveillance is surveillance [13]. However, these Infodemiology and infoveillance helps in measuring public attention, attitudes, behaviour, knowledge, and information consumption, as well as for syndromic surveillance, health communication, and knowledge translation research [14]. Moreover, the infoveillance data plays a crucial role to increase awareness, understand the disease transmission pattern, implementation of appropriate interventions and developing public health policy [15-17]. The Infodemiology and infoveillance approaches provided valuable information for monitoring, and forecasting outbreaks such as Ebola, Zika, MERS, influenza, measles, dengue, HIV/AIDS, erythromelalgia and syphilis [18-27].

The exploitation of internet data as a source to characterize epidemiological patterns for communicable and non-communicable diseases has been using since the mid-90’s under the concept of digital epidemiology. Digital epidemiology provides a valuable support for traditional epidemiology, where it can contribute to identifying emerging human pathogens, outbreaks of infectious diseases, epidemics even earlier than using the traditional epidemiological approach [28]. Google Trends (GT) is one of such innovative and efficient trend analyser to assess the internet search patterns and it tested as a useful tool for tracking social trends. GT provides big data based on geospatial and temporal pattern analysis focused on search queries of the public at different time points conducted through Google. The search results can filter geographically, temporally, categorically, and type of search. In the area of infectious diseases, GT provides valuable information and fascinating insights of health related issues, patterns of diseases at the community level [29,30]. A feature in GT, which allows users to plot the frequency of searches for a term, a string of multiple terms, or a phrase and also assess the traffic for the selected term and normalized on a relative, rather than absolute, basis [31]. GT was a proven tool for assessing malaria prevalence in countries with low levels of internet facilities [32]. Similarly, a high correlation was observed between the web search queries and the percentage of patients with influenza-like symptoms at a specific point of time. Moreover, GT can detect influenza transmission in 1 or 2 weeks earlier than the CDC [33]. The Google search queries are accurate and effective for predicting dengue incidence in countries (Bolivia, Brazil, India, Indonesia, Singapore and Mexico) which have poor surveillance systems [13,15,16].

Recent studies have shown that the GT data could be useful for tracking the COVID-19 cases at the population level as well as a significant correlation observed between search keywords and COVID-19 cases [34-36]. Unlike these studies, the present study has focused on using internet search data for tracking pandemic spread as well as to draw the relation between clinical conditions searched by the population and the number of daily COVID-19 cases in India and few selected states of the country. Similarly, we examine the time-lag correlation analysis between searched clinical course and reported COVID-19 case count. Followed by this, the study focuses on the analyses and predicting the COVID-19 cases using the Bayesian regression and linear regression models.

## 2. Methods

Here, figure-1 shows the overall workflow of the study. The whole study was classified into three parts, which are (1) data collection (2) data pre-processing and (3) data analysis.

**Fig-1.**
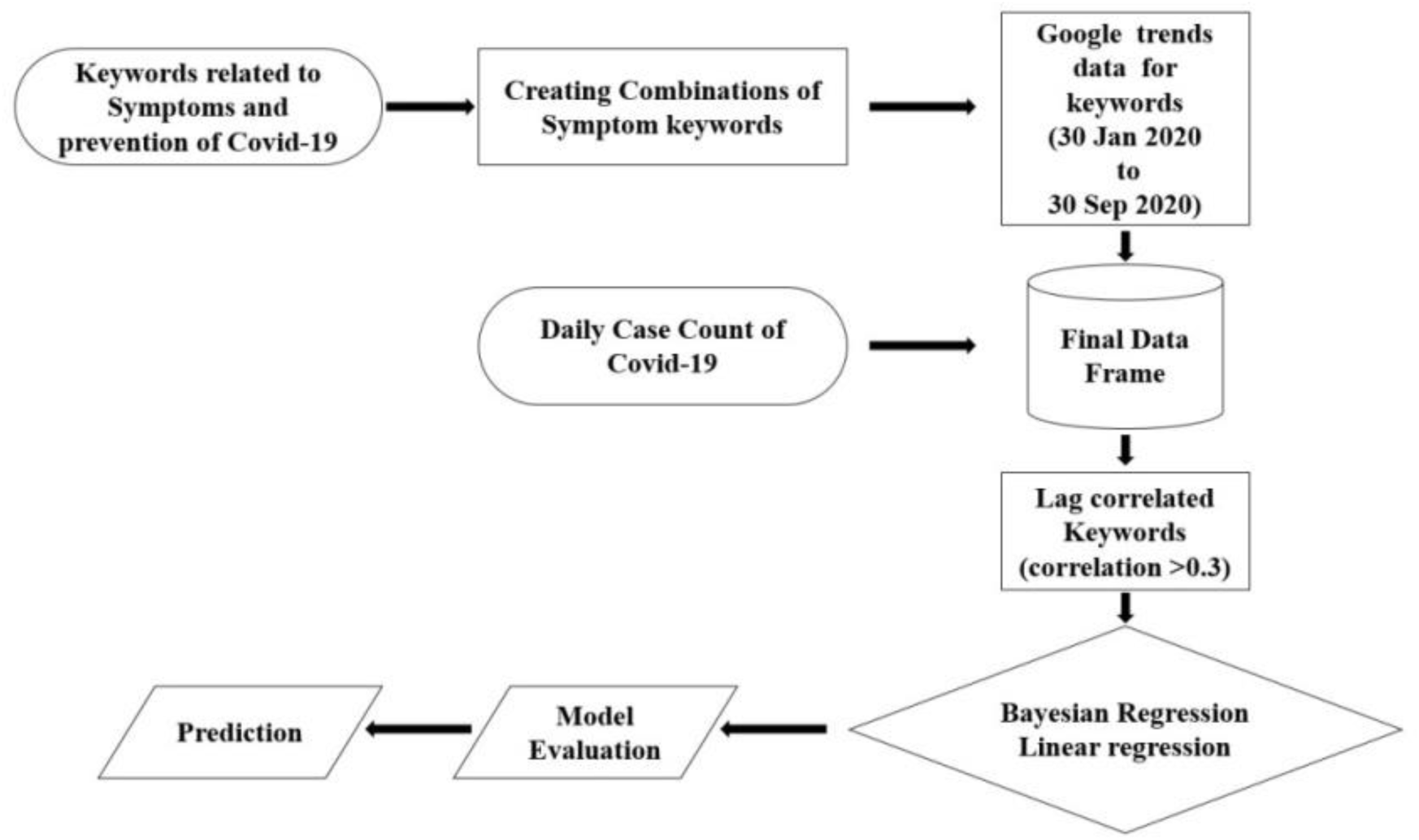
Overall work flow the study

### 2.1. Data collection

#### Google Trends Data

GT provides an index of the relative volume of search queries conducted through Google. GT symbolizes interest over time as the relative search volume (RSV) for the given region and time, wherein the highest peak period within the queried timeframe is reported as RSV = 100 (Google 2020). The search volume in GT is the traffic for a precise search term relative to all enquiries made in Google, normalized to range from 0 to 100. A value of 100 is the search term is popular; a value of 50 means the search term is partially popular. Similarly, a score of ‘0’ means there was not enough data for this search term. GT provides the RSV on a daily or weekly basis depending on the duration of the queried timeframe [37].

Search terms from GT related to various clinical symptoms of COVID-19 were retrieved for the nation India and few selected states from the Google Trends website [https://trends.google.com/trends] from January 30, 2020, to September 30, 2020. The retrieved Google Trends search index ranges from 0 to 100. The keywords used in the search volume index are "cold, headache, loss of taste, cough, fever, difficulty in breathing, loss of smell, nasal congestion, runny nose, sore throat, sneezing, tiredness, diarrhea, hand rub, hand wash, hand sanitizer, alcohol sanitizer, mask, touching eyes, nearest hospital, respiratory hygiene and, cover face". An explicit set of all possible combinations (up to 3 terms) of symptoms keywords were created. A final list of keywords made up of 386 keywords among which 13 were with the single search term of symptoms, 364 are a combination of symptoms search terms and nine keywords consist of preventive measure (Table-S1). However, GT search index data detected only 81 keywords across India. Finally, the Google Trends index summarized all search terms of clinical symptoms and preventive measures (81 keywords) of COVID-19 to build a data set. GT data were then compared with daily data on COVID-19 cases.

#### COVID-19 Case Data

All 28 states and 08 Union Territories of India covering latitude (8°N-38°N) and longitude (68°E-98°E) considered for the study. Daily counts of laboratory confirmed COVID-19 cases of India collected from the Ministry of Health and Family Welfare (MoHFW, https://www.mohfw.gov.in/), Government of India (GOI) from January 30, 2020 to September 30, 2020, [38].

### 2.2. Data pre-processing

To remove inherent variability in the data, standardization is a common approach in many machine-learning estimators for putting different variables on the same scale. Standardization scales of each input variable separated by subtracting the mean (called centering) and divided by the standard deviation to shift the distribution to have a mean of zero and a standard deviation of one for all variables. These standardization leads are flexible for comparison of the impact of the explanatory variables on the dependent variable. This preprocessing analysis was carried out by using the python library *sklearn.preprocessing.StandardScaler* [39].

### 2.3. Data analysis

Pearson’s correlation analysis was performed between search keywords of coronavirus symptoms (RSVs) and daily COVID-19 cases. Similarly, to assess the temporal relationship, the cross-correlation analysis was performed to calculate the time lag effect between RSVs and daily COVID-19 cases in India. This analysis was performed by using python library *statsmodels.tsa.stattools.ccf* [40]. Statistical significance was set as p<0.05. Rather than the direct correlation, the time lagged cross correlation showed a strong association between RSVs and COVID-19 cases. The explanatory variables, which were having absolute cross correlation coefficients greater than 0.3 with confirmed cases were considered for further analysis. The final dataset was split into two datasets i.e. train set with 75% values and test set with 25% values. Followed by, a COVID-19 predictability analysis based on Google Trends time series for India and few states was performed using Bayesian Regression along with its frequentist counterpart, linear regression.

#### Linear Regression

The basic linear regression in the frequentist approach holds the assumption about prediction as follows:

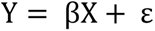

To generate response *Y*, the independent variables gain individual weights (**β′*s***) along with some noise due to random sampling. In the ordinary least squares (OLS) method, weights get values by minimizing the sum of squared errors on the training data. These values are a single point to estimate the model weights, also called parameters in OLS method. Finally, these point estimates used to make predictions for new data points.

#### Bayesian Linear Regression

Bayesian linear regression is an approach to linear regression in which the statistical analysis carried out in the Bayesian inference. There is a prior, likelihood distribution and posterior distribution are the three major steps in the Bayesian approach [41]. Bayesian Linear Regression using a probability distribution like normal (Gaussian) distribution, which can be expressed as:

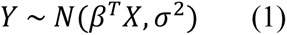

The model parameters assumed to be drawn from a probability distribution in Bayesian Models [42]. In Eq. (1), the response Y is generated from the Gaussian distribution with mean equals to the product of input variable values, model parameters and a standard deviation σ. The Bayesian Linear Regression aims to determine the posterior probability distribution for the model parameters given the inputs, X, and outputs, y:

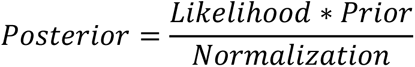

i.e.

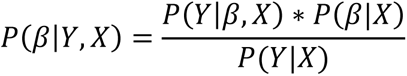

Here ***P*(*β*|*Y*, *X*)** is the posterior distribution, ***P*(*Y*|*β*, *X*)** is the likelihood of the data, ***P*(*β*|*X*)** is the prior and a normalization constant ***P*(*Y*|*X*)**. This is a simple expression of Bayes theorem.

The choice for prior is completely based on the domain knowledge, which means that we can add our prior knowledge of how data has been revolving around, through probability distributions. Thus, we can also able to quantify the uncertainty in parameters, unlike linear regression. A common prior choice is to use a normal distribution for β and a half-Cauchy distribution for σ. The computation of posterior distribution is extremely expensive and the sampling method Markov Chain Monte Carlo (MCMC) to approximate the posterior for each of the model parameters [43]. Monte Carlo methods are a class of computational algorithms, which involves drawing random samples from a probability distribution, where in Markov Chain controls the flow of random sampling in such a way that the next sample draw completely depends on its previous draw [44]. In Python PyMC3 package was used for Bayesian, linear Regression, which focuses on advanced Markov Chain Monte Carlo and variance fitting algorithms [42].

The outcome of the Bayesian Linear Regression is not a single estimate of the model parameter; however, the prediction of new observations drawn based on posterior distributions. Similarly, the mean of posterior distributions of model weights multiplied with new data points for prediction of cases (equation-1).

### 2.4. Model evaluation metrics

To evaluate the models, Mean Absolute Error (MAE) and Root Mean Squared Error (RMSE) used as evaluation metrics:

The Mean Absolute Error (MAE) between the predicted values 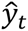 and the true values *y*_*t*_ can be calculated by using the formula mentioned below:

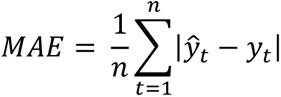

The Root Mean Square Error (RMSE) between the predicted values 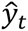 and the true values *y*_*t*_ can be calculated by using the formula mentioned below:

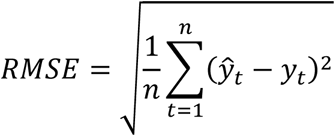

### 2.5. Statistical Software

Python 3.7.4 and its packages such as PyMC3 3.8, statsmodels 0.12.2, pandas 0.25.1, numpy 1.18.5, scikit -learn 0.21.3, matplotlib 3.2.2.

## 3. Results

### 3.1. COVID-19 in India

The first case of COVID-19 infection reported in Kerala, India on 30 January 2020 as the infected person returned from Wuhan, China [45]. In the following weeks, the number of COVID-19 cases as well as deaths increased steadily that are associated with travel activities. To reduce the spread of disease transmission, the Government of India has imposed a lockdown from 25 March to 31 May 2020 [46]. As of November 18, 2022 India, is the second most COVID-19 infected country in the World and the first in Asia witnessed 4,46,67,967 confirmed cases with 7,034 (0.01%) active cases, along with 4,41,30,380 recovered cases (recovery rate of 98.79%) followed by 5,30,553 deaths. Figure-2 shows the daily new cases and deaths of COVID-19 in India from January 30, 2020 to November 18, 2022. Figure-3, illustrate the spatiotemporal distribution of COVID-19 prevalence by state wise in India as of November 18, 2022. COVID-19 is encountered in all states but, half of the disease burden is contributed by the states such as Maharashtra (81,34,891[18.2%]), Kerala (68,24,839[15.2%]), Karnataka (40,70,642[9.1%]), Tamil Nadu (35,93,713[8.0%]) and Andhra Pradesh (23,39,039[5.2%]) [38]. However, Andaman and Nicobar Islands (10,741), Lakshadweep (11,415), Dadra and Nagar Haveli and Daman and Diu (11,591), Ladakh (29,398), and Nagaland (35,986) have reported the least number of cases (Fig.3). Though there was a very limited number of tests for COVID-19 during the initial stage of the pandemic, and later the number of tests were gradually increased, and as of November 18, 2022, a total of 90,35,50,540 tests were conducted in India. Similarly, the world’s largest COVID-19 vaccination program is rolled out by Government of India on January 16, 2021 and a total of 219.85 crore vaccines were administered as of November 18, 2022. [47].

**Fig. 2:**
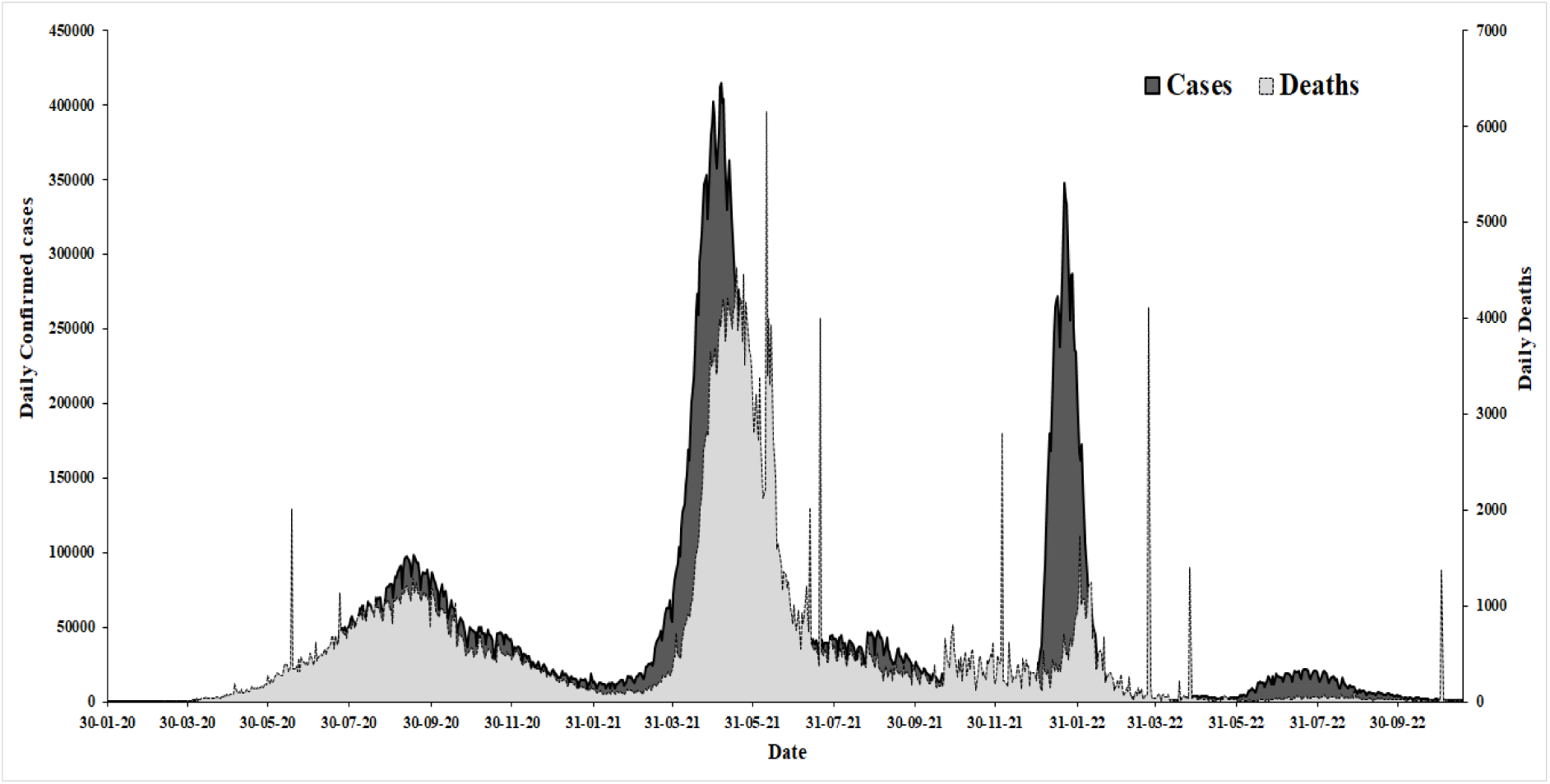
Day wise confirmed COVID-19 cases in India.

**Fig.3:**
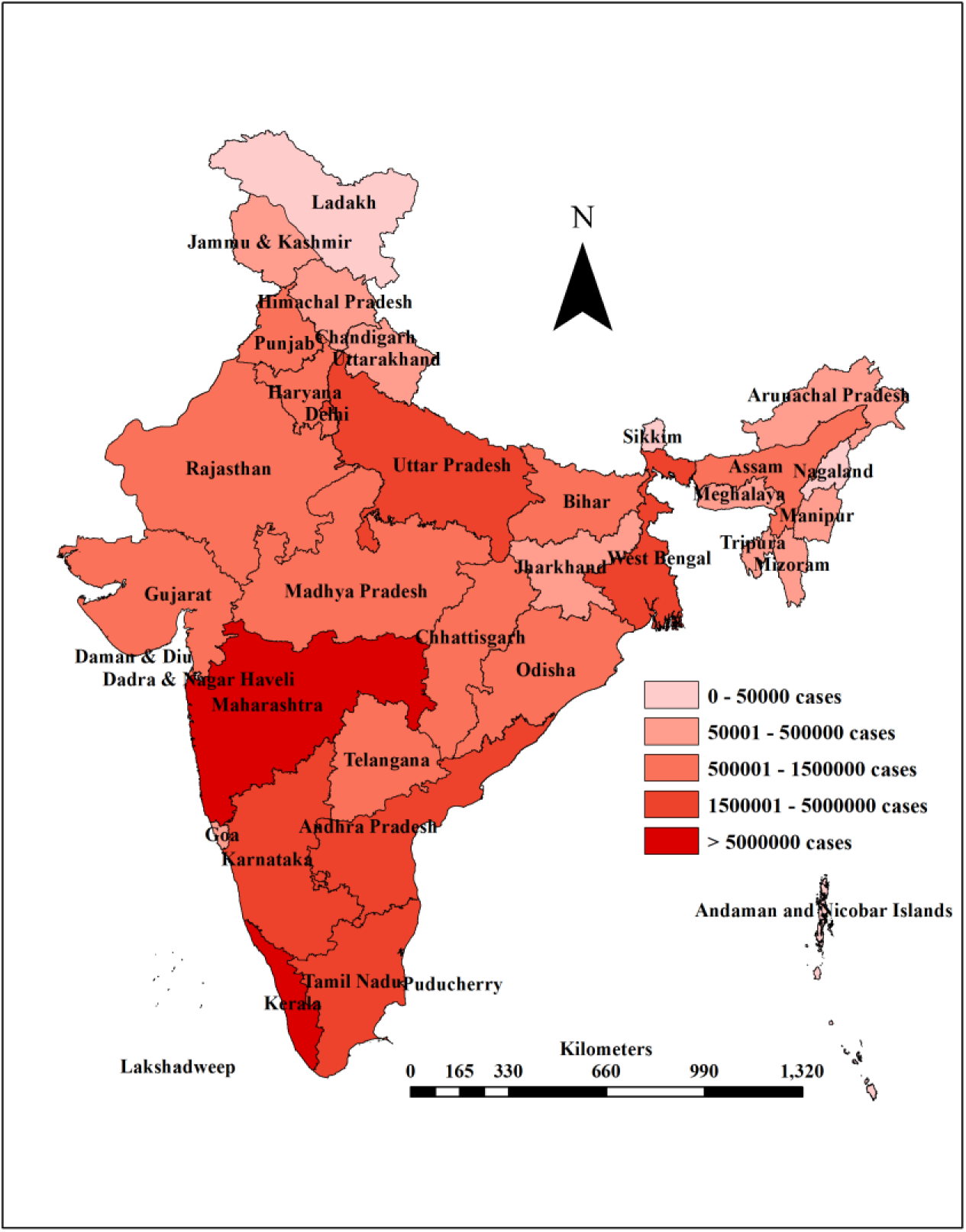
Spatial distribution of state wise COVID-19 cases in India.

Figure-4 shows the RSVs of selected keywords related to COVID-19 clinical symptoms and preventive measures such as "cold, headache, loss of taste, cough, fever, difficulty in breathing, loss of smell, nasal congestion, runny nose, sore throat, sneezing, tiredness, diarrhea, hand rub, hand wash, hand sanitizer, alcohol sanitizer, mask, touching eyes, nearest hospital, respiratory hygiene and, cover face" along with the daily COVID-19 cases across India during the first wave of COVID-19 pandemic.

**Fig.4:**
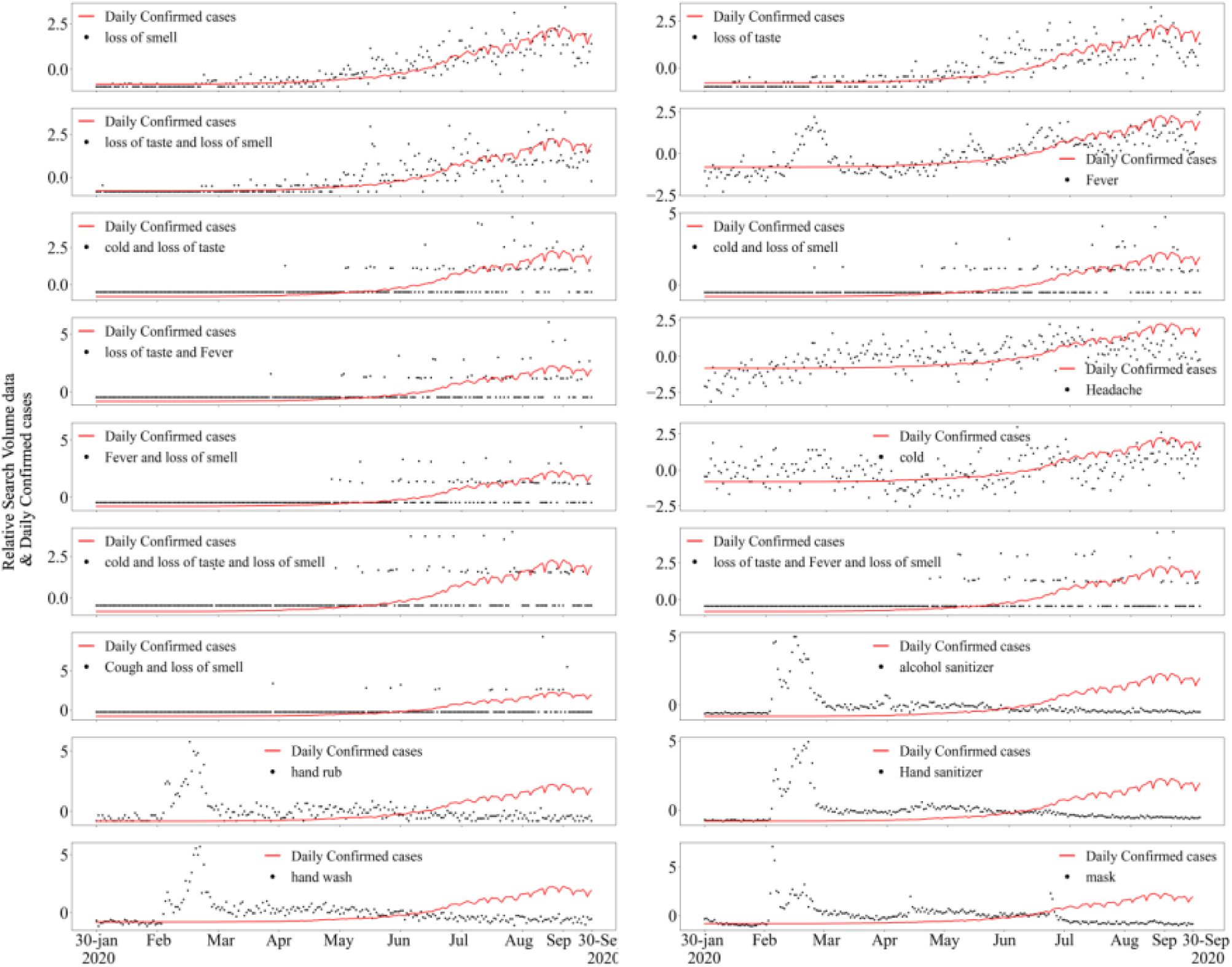
Scaled Google Trends data for the selected search keywords and daily-confirmed Covid-19 cases across India. The feature selected here based on correlation value (R=>0.3).

### 3.2. Correlation analysis

Correlation analysis was performed between each search term and daily reported new COVID-19 cases of India as a nation (Table-1). The correlations ranged from R=-0.02 (respiratory hygiene) to R=0.86 (loss of smell) with daily new COVID-19 cases. Among all search keywords cold and headache have shown moderate correlation (R=0.41 & 0.46), while fever, loss of taste and loss of smell had strong correlations (R=0.66, 0.78 & 0.86) with COVID-19 cases.

**Table-1:**
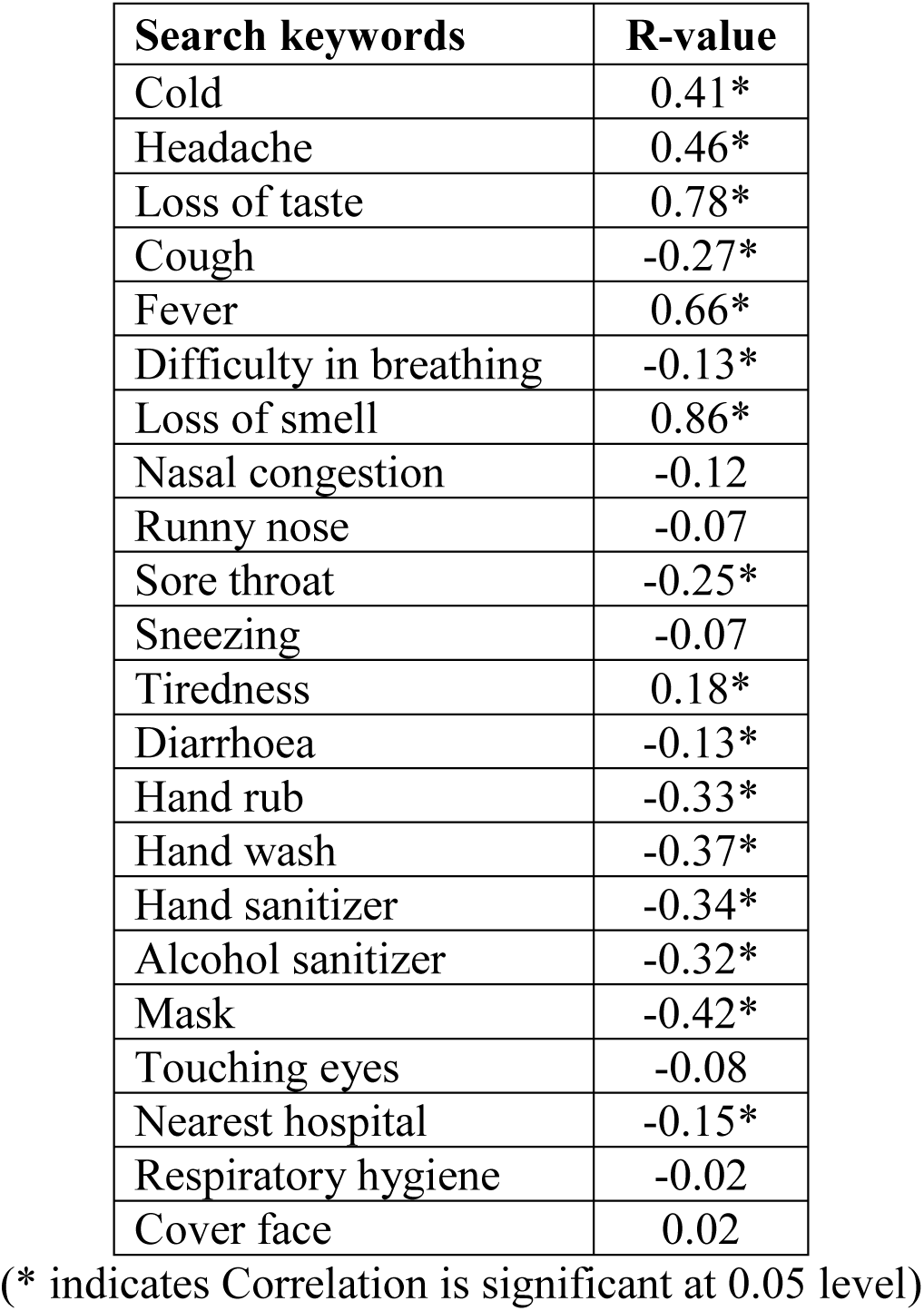
Correlation coefficient values between search terms and COVID-19 cases in India.

The cross-correlation analysis shows that most of the search keywords had moderate to strong correlations observed with time lag period (Figure-5). The keyword ’loss of smell’ shows a significant correlation (R=0.89) with COVID-19 cases with three days lag period. Similarly, ’loss of taste’ (R=0.82), ’loss of taste and loss of smell (R=0.76)’, ’fever’ (R=0.68), and ’headache’ (R=0.55) shown a significant correlation with COVID-19 cases with a lag period of 9, 1, 9 and 13 days respectively. The preventive measures such as ’mask’, ’hand wash’, ’hand sanitizer’, ’hand rub’, and ’alcohol sanitizer’ has shown a negative association with zero lag period (Fig.5). The lag period (days) and its correlation coefficient values for other search keywords shown in table-2.

**Fig.5:**
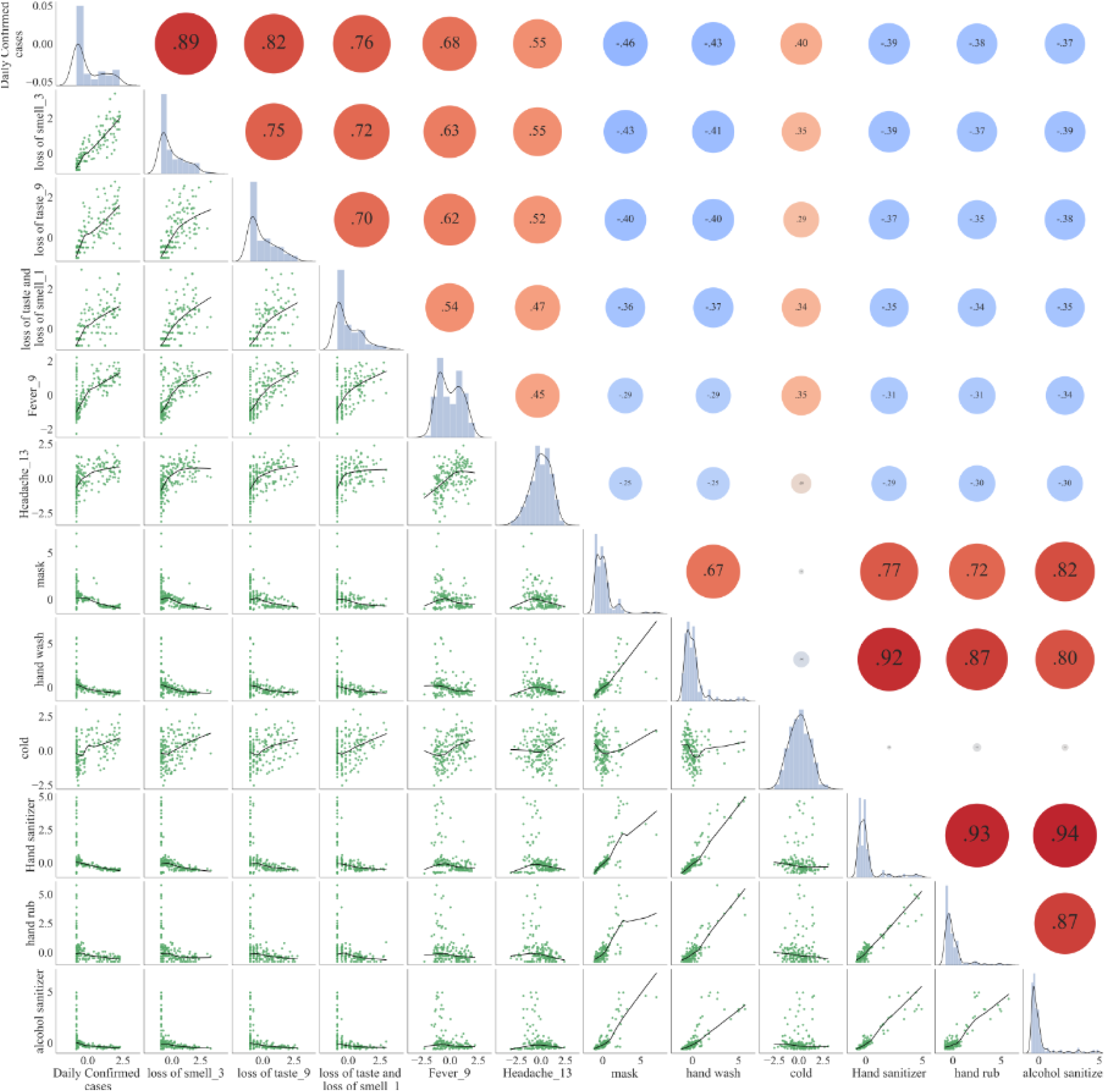
Time-lag correlations of search terms of clinical symptoms of COVID-19 and newly confirmed daily COVID-19 cases for the entire India. Out of 23 search keywords only 11 variables were selected based on correlation analysis and variables which have R>0.3.

**Table-2:** The time lag (days) and correlation coefficient between the search volume for specific search term and reported daily COVID-19 cases for whole India.

| Search word (RSVs) | Lag period | R-value |
| --- | --- | --- |
| Loss of smell | 3 | 0.87 |
| Loss of taste | 9 | 0.81 |
| Loss of taste and loss of smell | 1 | 0.76 |
| Fever | 9 | 0.68 |
| Headache | 13 | 0.55 |
| Cold | 0 | 0.42 |
| Cold and headache | 19 | 0.27 |
| Headache and fever | 14 | 0.19 |
| Fever and tiredness | 18 | 0.14 |
| Cold and sneezing | 3 | 0.14 |
| Headache and cough | 18 | 0.11 |
| Headache and sore throat | 7 | 0.11 |
| Cold and tiredness | 0 | 0.1 |
| Cold and diarrhoea | 0 | 0.06 |
| Cold and nasal congestion | 12 | -0.08 |
| Cold and fever and runny nose | 2 | -0.09 |
| Headache and diarrhoea | 9 | -0.11 |
| Cough and nasal congestion | 14 | -0.15 |
| Cough and fever and sore throat | 8 | -0.16 |
| Cough and sore throat | 10 | -0.25 |
| Sore throat | 0 | -0.25 |
| Mask | 0 | -0.36 |
| Hand wash | 0 | -0.36 |
| Hand sanitizer | 0 | -0.38 |
| Hand rub | 0 | -0.42 |
| Alcohol sanitizer | 0 | -0.47 |
| Tiredness | - | - |
| Fever and sore throat | - | - |
| Cold and sore throat | - | - |
| Respiratory hygiene | - | - |
| Cold and runny nose | - | - |
| Lung disease | - | - |

### 3.3. Bayesian Regression Analysis

As shown in figure-6, each variable is following a Gaussian distribution, hence, normal prior is considered for each of the independent variables in the analysis. The Bayesian regression model was built by using the training dataset. PyMC3 library in the python programming language is used for the study and it has automatically chosen No-U-Turn sampler to draw the posterior distribution. This sampler stores the samples for each of the model parameters in a trace. The trace plots are assessing convergence of individual parameters. The trace plots for all the variables for India is shown in figure-6. Trace plots provide the posterior distribution of each modelled parameter on left-hand side, and the horizontal band formed from the progression of the samples converged properly in the chains were shown on the right-hand side of the figure-6. The color lines in the plots represent the Markov chains (Fig-6).

**Fig.6:**
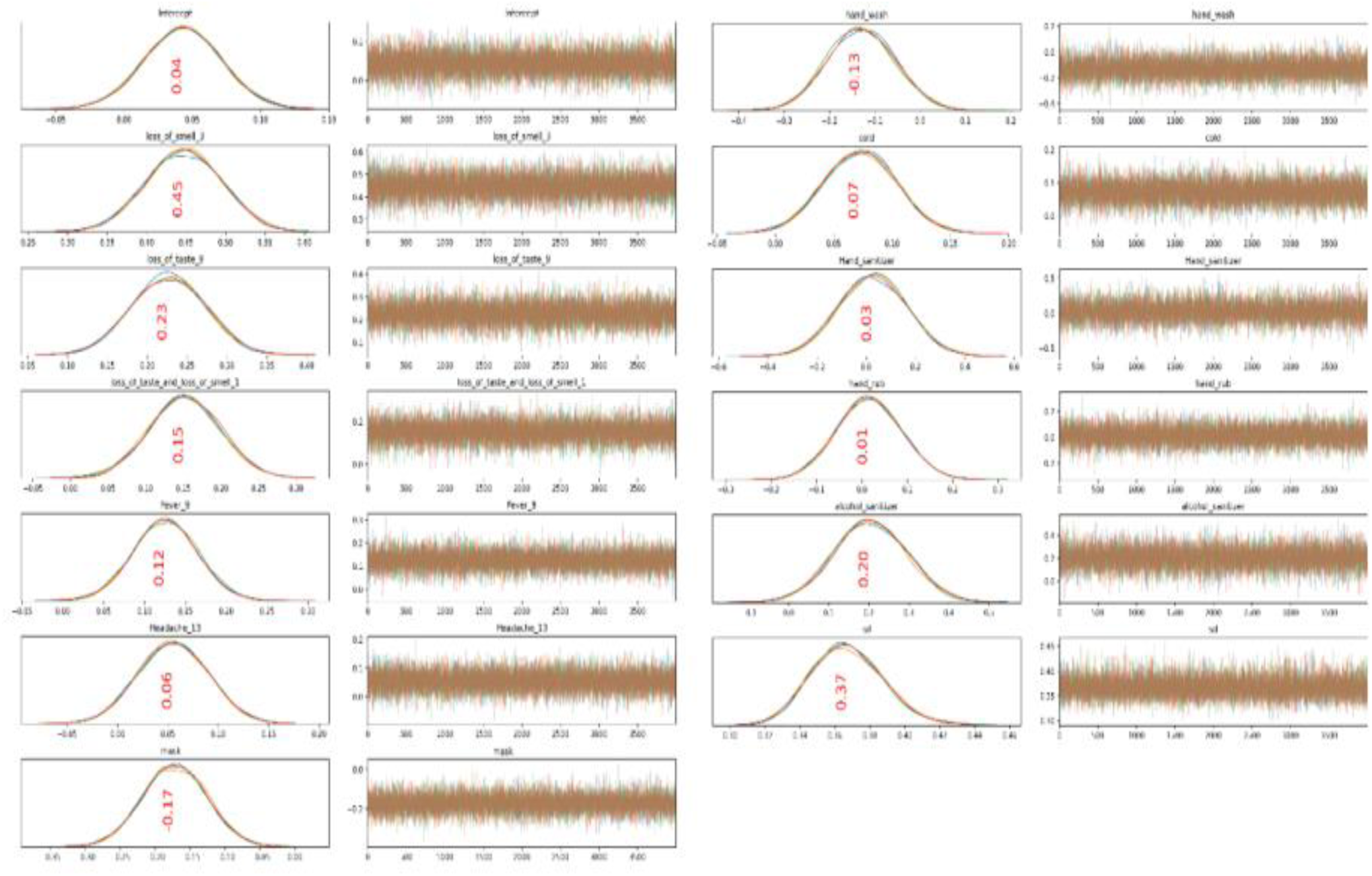
Trace plot for posterior distributions of each parameter and their progression of samples drawn for entire India.

Figure-7 illustrates the 94% Highest Posterior Density (HPD) which is the credible interval for RSVs parameters. The credible interval is an interval within which an unobserved parameter value maintains a given probability. The mean for 94% confidence intervals (CI) for various search keywords is also provided in the plot (Fig.7).

**Fig.7:**
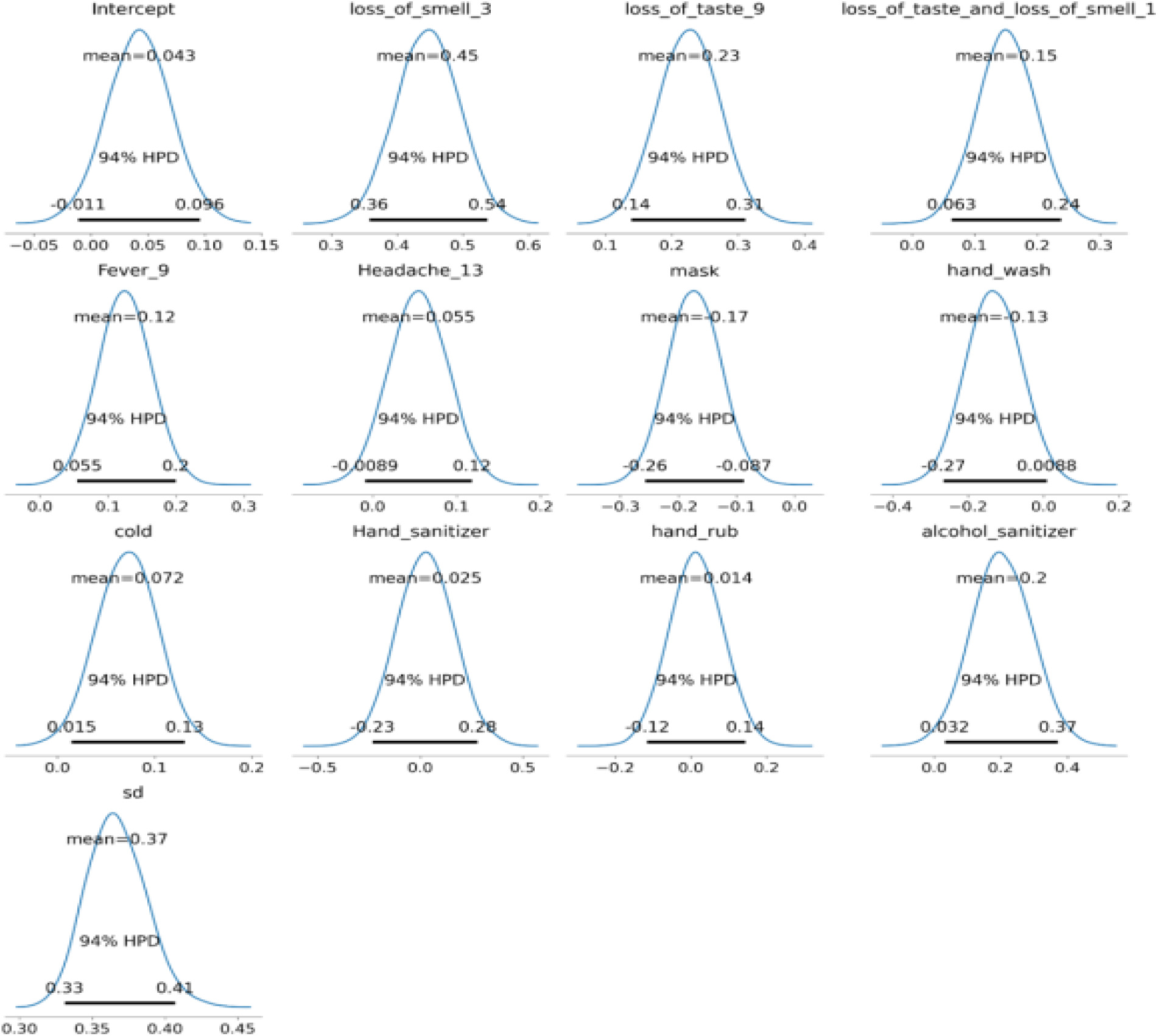
Highest Probability Density (HPD) plots of each parameter with their 94% credible intervals for India.

The posterior distribution for prediction of COVID-19 cases by using the means of 94% HPD of various parameters interpreted as:

Daily_Confirmed ∼ N (0.04 * Intercept + 0.45 * loss_of_smell_3 + 0.23 * loss_of_taste_9 + 0.15 * loss_of_taste_and_loss_of_smell_1 + 0.12 * Fever_9 + 0.05 * Headache_13 + -0.17 * mask + -0.13 * hand_wash + 0.07 * cold + 0.02 * Hand_sanitizer + 0.01 * hand_rub + 0.20 * alcohol_sanitizer + 0.37^2^).

Figure-8 explains the probable density plots of COVID-19 and the entire distribution. The predictions obtained by using the posterior distributions for two test data set observations (15^th^&25^th^) is shown in figure-8. The mean estimate of the predicted COVID-19 cases distributions is equal to the true mean estimate.

**Fig. 8:**
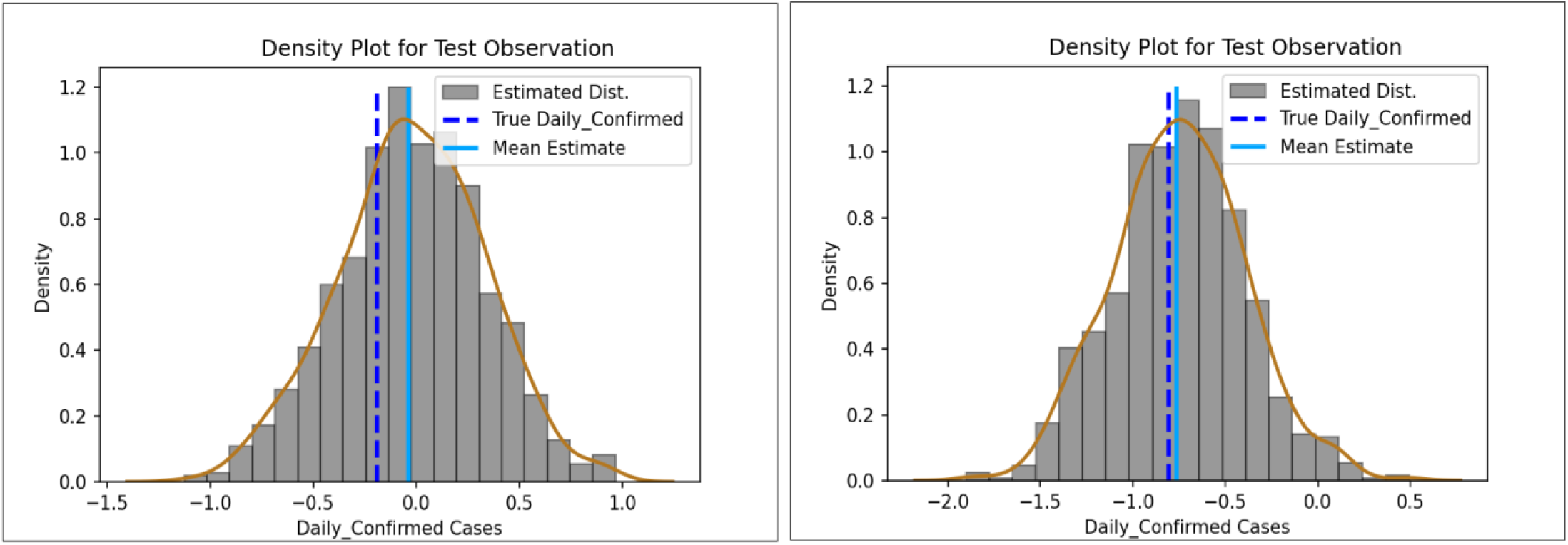
Observed and predicted COVID-19 probability density distributions for India.

Besides Bayesian regression, the classical linear regression model also carried out to predict the COVID-19 cases for the nation as well as for states Maharashtra, Andhra Pradesh, Tamil Nadu, Kerala and Delhi. Among both the models, the Bayesian regression model outperformed the frequentist linear regression. The model evaluation metrics such as mean absolute error (MAE) and root mean square error (RMSE) for both the models of different states were presented in table-3.

**Table-3:** The model evaluation metrics MAE and RMSE for India and different states.

| Location | Bayesian Regression |  | Linear Regression |  |
| --- | --- | --- | --- | --- |
|  | MAE | RMSE | MAE | RMSE |
| India | 0.331164 | 0.411087 | 0.33134 | 0.411316 |
| Maharashtra | 0.500642 | 0.629627 | 0.500829 | 0.629934 |
| Andhra Pradesh | 0.326427 | 0.527788 | 0.326432 | 0.527791 |
| Tamil Nadu | 0.618469 | 0.718733 | 0.61866 | 0.718687 |
| Kerala | 0.722255 | 1.04137 | 0.722661 | 1.04202 |
| Delhi | 0.547009 | 0.729039 | 0.547023 | 0.729019 |

## 4. Discussion

There is an urgent need for proactive identification and prediction of the spread of novel infectious diseases for public health importance, especially when unexpected epidemics/ pandemics occurs and which are not recorded by traditional surveillance systems [48]. In recent years, the number of studies has pointed out the significant role of Internet surveillance in helping the public health authorities for early prediction of disease outbreaks [18-27]. Since the past decade, internet-based search has gradually increased globally [49]. During the pandemic period, the Internet search patterns relating to coronavirus, corona and COVID-19 is very high and many researchers used these search data index from Google Trends for understanding the disease dynamics [30,36]. In contrast to the earlier studies, the present study used the search terms of clinical symptoms of COVID-19 GT data to draw the association with real-time COVID-19 cases and predicted the COVID-19 using Bayesian regression and linear regression across India and few states of the country.

However, the present study demonstrated that the data obtained from the Google Trends index on daily search keywords on clinical symptoms of COVID-19 has moderate to strong association with laboratory confirmed COVID-19 cases. We also found that the peak search for these keywords in Internet search engines was 10–14 days earlier than the incidence peak of COVID-19 observed [50]. The lag correlation showed a minimum and maximum correlation at 2-19 days observed. However, previous studies have reported that the maximum lag period for predicting COVID-19 cases was 21 days in India [36]. After the initial cases of COVID-19 in Kerala, the number of cases is gradually increasing in various parts of India. To flatten the COVID-19 curve, India suspended visas for all international travelers from March 13, 2020, onwards. Followed by a travel ban, the Government of India announced a nationwide lockdown (from March 25 to May 31, 2020) to minimize human activity across the country [51]. Similarly, COVID-19 testing capability increased rapidly to identify and isolate the infected populace for minimizing the diseases spread [52].

The infoveillance (i.e. surveillance) approach is the passive method, here the data is collected based on internet search trends passively without involving individuals [13]. In recent years Google Trends and Twitter has identified as better data sources for infodemiology (i.e. epidemiology) while other social media platforms such as Facebook and Instagram help to understand the online behavioural pattern like level of understanding the health issues, knowledge about health-related conditions [13,16]. To the best of the author knowledge, this study is the first to conduct the COVID-19 symptoms related search term for India. The study used different search terms like single keyword (e.g. cough), double keywords (e.g. cough + cold) and triple keywords (e.g. cough + cold + fever). This kind of search provides accurate and reliable information for understanding the clinical course of illness for COVID-19. Similarly, it also helps the hospitals, public health care workers to assess the disease burden and improve the health care facilities to patients.

The goal of the study is to assess the internet based infoveillance survey used as a complementary data survey during the novel pandemics for understanding the clinical course of the disease. In this line, the GT data is used for qualitative analysis for prediction of COVID-19 in India as well as state level in Maharashtra, Andhra Pradesh, Tamil Nadu, Kerala and Delhi. Considering this data, Bayesian regression performed better than the linear regression and provide very promising results in predicting the COVID-19 at the Indian and regional level. This contribution supplements the public health authorities to implement appropriate control measures to handle the spread of the disease.

## Conclusions

The web-based search activity plays a vital role in recognising the early detection of infectious disease outbreaks, further, it helps in preparing the healthcare facilities and to implement appropriate control activities timely to avoid adverse impacts. The present study reveals the use of internet surveillance and makes use of Google Trends to monitor new infectious diseases such as COVID-19 and to develop predictive models to understand the transmission dynamics of the disease. The GT data showed a moderate to strong association with COVID-19 cases across the India. There is an important finding of a consistent pattern between GT data and COVID-19 incidence at the state level (Fig.S1 for Maharashtra state). The predictability of Bayesian regression analysis provides insights of online search data that can play an important role in forming public health policies, especially in times of epidemics and outbreaks.

## Supporting information

Fig.S1 for Maharashtra state

## Data Availability

All data produced in the present study are available upon reasonable request to the authors

## Acknowledgments

The authors are grateful to the Director of the Council of Scientific and Industrial Research-Indian Institute of Chemical Technology, Hyderabad, for his encouragement and support. The present work is supported by the DST (Department of Science and Technology) under Epidemiology Data Analytics (EDA) of Interdisciplinary cyber-physical systems (ICPS) programme (Grant number: DST/ICPS/EDA/2018), Govt. of India and the Ministry of Environment, Forest & Climate Change (MoEF & CC), Government of India for funding the project Environmental Information Awareness Capacity Building and Livelihood Programme (EIACP: Resource Partner on Climate Change and Public Health). The funders had no role in study design, data collection, and analysis, decision to publish, or preparation of the manuscript. CSIR-IICT communication number of the article is IICT/Pubs./2021/074.

## Ethical Statement

The authors declare that an ethical statement is not applicable because the case information has been gathered.

## Data Availability Statement

The data used in this study are available from the corresponding author upon request.

## Authors Contributions

Hariprasad Vavilala: Data curation; Methodology; Formal analysis; Validation; Visualization; Writing - original draft. Rajasekhar Mopuri: Spatial mapping and data collection and processing. Srinivasa Rao Mutheneni: Conceptualization; Formal analysis; Investigation; Methodology; Project administration; Supervision; Writing-review & editing.

## Supporting Information

Fig-S1: Scaled Google Trends data for the selected keywords and Daily confirmed Covid-19 Cases of Maharashtra.

Table-S1: A final list of search keywords and a combination of keywords for COVID-19.

