## Supplementary figures and images for "Assessment of relationship between Google Trend search data on clinical symptoms and cases reported during the first wave of COVID-19 outbreak in India"

### Fig.S1 for Maharashtra state

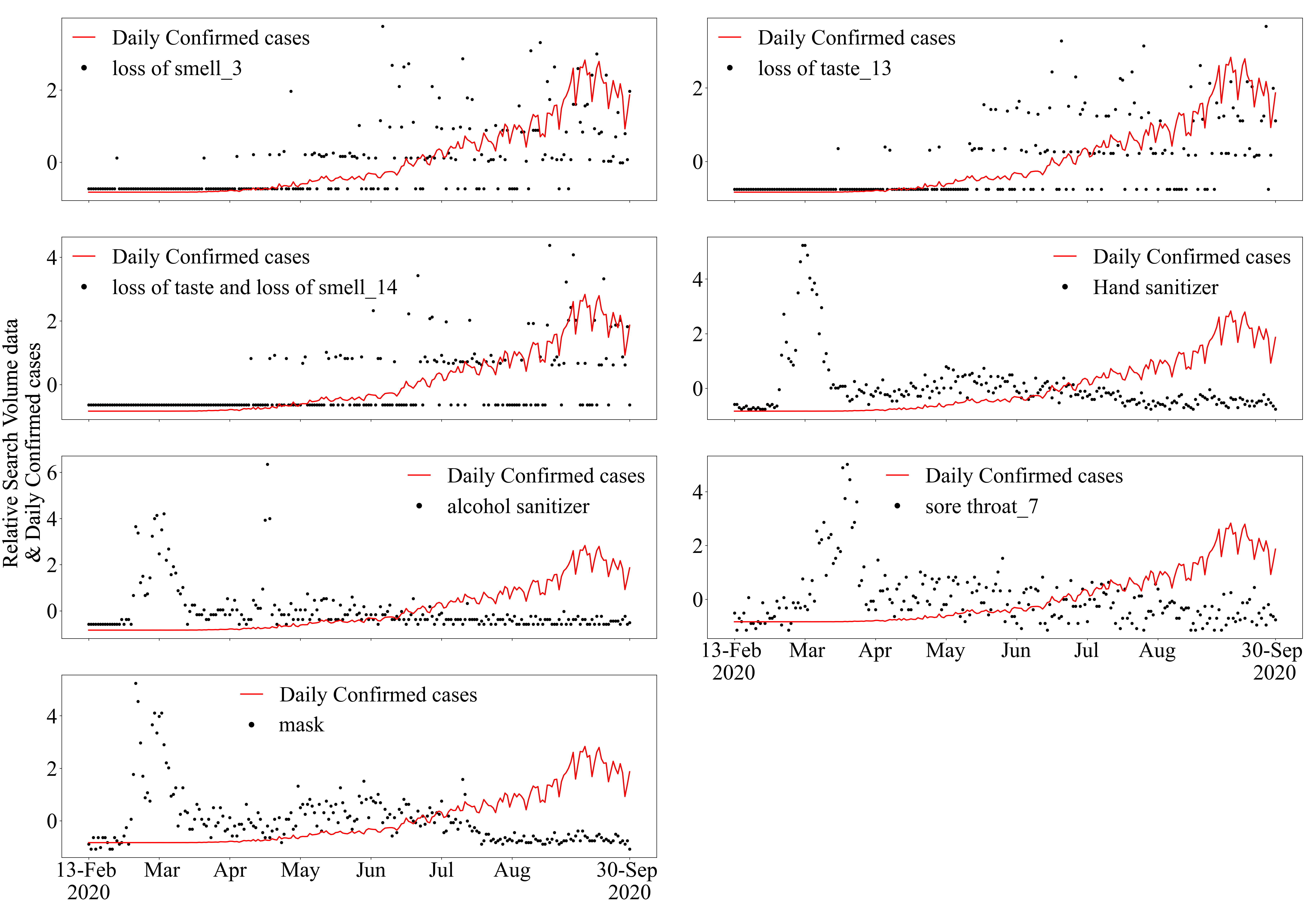
